## Supplemental Information for "Minding the margins: Evaluating the impact of COVID-19 among Latinx and Black communities with optimal qualitative serological assessment tools"

### Blood Collection/Processing and On-site Testing

Up to 500 μL of blood was collected with Tasso SST devices (Tasso, Inc., Seattle, WA) as as per manufacturer's guidelines. Briefly, the device was attached to the participant's arm and a button released a small needle upon pressing, the pre-attached tube (containing no-anticoagulants or other additives) then filled with blood within 5-10 minutes.

The blood-filled tube was detached, and one drop of whole blood used for the US Food and Drug Administration (FDA) emergency use authorized (EUA) approved point of care test (FaStep from Assure Tech, Hangzhou, China) to measure SARS-CoV-2 anti-immunoglobulin (Ig)G and IgM antibodies as indicated by the manufacturer. The FaStep test is a rapid lateral flow chromatographic immunoassay for qualitative detection and differentiation of IgM and IgG antibodies to SARS-CoV-2 Spike and Nucleocapsid. The POC test results were communicated back to the participants immediately, explaining that the outcomes should not be used to make any clinical decisions or behavioral changes regarding COVID-19 transmission prevention measures. Further, participants were offered a $50 time and travel reimbursement in the form of a Visa gift card.

The rest of the blood was transported to the laboratory, incubated at room temperature (RT) for 30 minutes, and centrifuged at 1,500 g for 10 minutes at RT to separate the serum. The serum fraction was aliquoted and stored at -20 °C for further serological screening, see Luminex Methods.

### Saliva Collection and Processing

Saliva samples were collected with SuperSal2 devices (Oasis Diagnostics, Vancouver, WA) as indicated by the manufacturer. Briefly, the collection device was held under the tongue of the participant until the red circle turned red, indicating saturation. The saliva was then squeezed out into a provided tube. Saliva samples were centrifuged at 10,000 g for 5 minutes at RT, the supernatant transferred to new tubes (removing debris) and stored at -20 °C for further serological screening, see Luminex Methods.

Banked saliva samples, collected with the same SuperSal2 devices, served as alternative saliva control group for the seroprevalence calculation. We obtained 50 banked adult saliva samples that were collected in September of 2020 at Ahero, Kisumu County, Kenya. The first laboratory-confirmed COVID-19 case in Kisumu County, Kenya was identified on June 9, 2020, and the first wave peaked in July 2020.(1)

### Multiplex Luminex Assay - Blood

The following SARS-CoV-2 antigens were coupled to Luminex MagPlex Microspheres as indicated by the manufacturer (see **Supplemental Table 5** for protein source information): Wild-type full-length spike, nucleocapsid, receptor-binding domain (RBD) Wuhan, RBD alpha, RBD beta, RBD gamma, RBD delta, RBD lambda, and RBD omicron. Additional human coronavirus (hCoV) and a Bovine Serum Albumin (BSA) control were coupled: hCoV HKU1, hCoV OC43, hCoV NL63, hCoV 229E.

After validation of conjugated beads, the participant blood samples were screened on 96- or 384-well plates following the Luminex Multiplex Immunoassay protocols on FlexMap3D equipment. Briefly, beads conjugated with each of the 20 antigens were combined and diluted in ABE buffer (PBS, BSA 0.1 %, TWEEN 20 %, Sodium Azide 0.05 %) and added to each well of a 96- or 384-well (500 beads per region/antigen for the panel bead mix). The plate was incubated on a magnet to fix the beads and the ABE buffer removed. Then 50 μL of serum samples diluted in ABE buffer (1:100) were added to the wells and incubated for 2 hours at RT on a plate shaker (300 rpm). Each plate also included a seven- or ten-point serial dilution of a prescreened positive control for SARS-CoV-2 (blood samples of confirmed COVID-19 cases). Following the incubation beads were washed three times with ABE buffer, post-magnet incubation. Then 50 μL of biotinylated anti-human secondary IgG or IgA (BD Pharmingen) diluted in ABE (1:1000) was added to each well containing the beads and incubated for 1 hour at RT on a plate shaker (300 rpm). Following incubation, beads were washed again three times with ABE and 50 μL of ABE diluted phycoerythrin conjugated streptavidin (1:1000; BD Pharmingen catalog # 554061) added to the wells containing the beads. After a final 15-minute incubation at RT on a plate shaker (300 rpm), a set of three ABE washes were performed and the beads resuspended in ABE (125 μL for 96-well plate and 75 μL for the 384 plate). After resuspension the plate was read on the FlexMap3D Luminex instrument where the median fluorescence intensity (MFI) of each antigen and bead count was recorded. Once the control MFIs, standards, and bead counts (minimum 50 beads per antigen in each well) were validated, BSA was subtracted (including for the standards) to account for non-specific binding. Previous studies showed that non-specific antibody binding (directly binding to the beads) may occur(2), which can be accounted for by subtracting the BSA-linked reads.

Previously described de-identified banked blood/serum samples served as negative blood controls (total n=50).[6] Briefly, SARS-CoV-2 negative banked blood samples were sourced from UMass Chan (collected between October of 2003 and January of 2020, n=19) and Case Western Reserve University (pre-screened samples from community blood drive, collected in July 2020, n=31). Previously described banked blood samples from COVID-19 patients hospitalized at UMass Memorial Hospital (collected between April and August of 2020) served as positive blood controls (n=50).[6]

Due to lack of adequate controls for the SARS-CoV-2 variants (RBD alpha, beta, gamma, delta, lambda, and omicron) and hCoVs (OC43, HKU1, 229E, and NL63) the serological outcomes (raw MFI minus BSA) for serum IgG were evaluated solely as quantitative results and not translated into qualitative outcomes.

**Multiplex Luminex Assay - Saliva**

To screen saliva for anti-SARS-CoV-2 and hCoV IgA and IgG antibodies the multiplex panel and Luminex methods were applied as described for blood, except that 50 μL of undiluted human saliva samples was added instead of serum. Additionally, the undiluted saliva samples were screened for total IgG and total IgA to account for differential salivation flow rates and therefore total antibody dilution in saliva by coupling anti-human IgG gamma chain (Bio-Rad, Hercules, CA) and anti-human IgA alpha chain protein (Abcam, Cambridge, UK) respectively, to Luminex MagPlex Microspheres as indicated by the manufacturer.

**Across-plate Normalization**

For the across plate normalization, dilution series of post-BSA subtracted standards for each antigen were weighted and a normalization factor determined and applied, rescaling the data to a weighted average scale.

Briefly, Luminex-based multiplex antibody measurements (MFI) quantify the concentration of antibodies (*c*). We assume that $log_2 c$ for dilution series saturates asymptotically and has a sigmoidal shape as a function of antibody concentration, which goes inversely as the dilution factor. Hence, concentration and dilution are interchangeable. The following dominant within-plate and across plate effects are considered here in order to obtain an across-plate normalization protocol. Within plate: serial dilution protocols lead to a compounding of uncertainties accumulated during pipetting (i.e., if the dilution was incorrect at an earlier point due to pipetting error, loss of liquid clinging to the pipetting tip, etc, the later dilutions will be incorrect as well). Accounting for this effect, we define the variance of the linear fit to be directly proportional to the dilution as detailed in the mathematical description below. Across-plate: The measurements performed on different plates could have high variance due to plate-specific factors such as background light during measurements, varying bead batches, instrument variations, among other factors. To account for this effect, we estimate the plate-specific variance from the data for that plate and weigh the plate in an inverse proportion to this variance in order to obtain an average plate, see below.

For a given antigen, we first determine the linear regime for the sigmoid curves corresponding to dilution curves post-BSA subtraction for all the plates and restrict the domain to this regime. Let $x_i$ be the dilution at step i, and let $y_{ij}$ be the $\log_2$ of the corresponding measurement value in this linear regime. We propose a dilution-dependent Normal distribution for each plate, with linear mean and variance proportional to dilution to account for compound uncertainties with increasing dilution. The proportionality constant here, $\sigma^2_j$, denotes the plate-dependent variance. That is, $y_{ij}$ is normally distributed as follows-

$y_{ij} \sim N(\, m_j x_i + s_j,\ i\sigma^2_j)$, for dilution I in {1,2,….,n} and plate j in {1,2,….,m}.

We then use the data available to define maximum likelihood estimator for each plate to find dilution-weighted best-fit lines using custom MATLAB fits, along the way estimating $m_j$, $s_j$, and $\sigma^2_j$. The slope and the intercept of the best fit line ($m_j$ and $s_j$) quantify the change in signal per unit change in dilution. The proportionality constant for the variance for each plate ($\sigma^2_j$) ascertains how well the data fit the line, i.e., it is a proxy of how linear the plate of interest behaves in this regime.

Hence, to normalize across plates, a reference standard dilution series was determined by averaging the standards of all plates encompassing one antigen/isotype combination by giving standards with less variance more weight. This way plates with less linear outcomes skewed the reference curve less. That is, the reference standard dilution curve has a value $z_i$ at the dilution $x_i$ as defined below-

$$z_i = \frac{1}{K} \Sigma_j \frac{1}{\sigma_j^2} \left(m_j x_i + s_j\right), where K = \Sigma_k \frac{1}{\sigma_j^2}.$$

Once the weighted average dilution curve was determined, the plate standards were translated to this curve to get a normalization factor (i.e., a single value for each antigen-plate pair) as follows- $L_j = \frac{1}{n} \Sigma_i \left(z_j - m_j x_i - s_j\right)$are the translation factors in $\log_2$-space, used to define $NF_j = 2^{L_j}$as the normalization factors for the given antigen for plate j.

Once the normalization factors were calculated, we applied it to the sample data by multiplying each antigen/isotype-specific sample MFI by the normalization factor corresponding to the plate. The transformed data was then pooled and used to determine the qualitative outcomes, see below.

HKU1 had low values across the standards (note the standards were chosen to target SARS-CoV-2) for IgA in serum and saliva. As BSA and these standard measurements were the same order of magnitude, post-BSA subtraction the values for these were within very small and often negative. Hence, we used the raw MFI measurements (without BSA subtraction) for all standards on the plates covering saliva and serum IgA for HKU1 to perform the across-plate normalization calculations. We additionally imposed the condition $m_j \leq 0$ to account for the fact that the signal should decrease with increasing dilution.

### Qualitative Outcome Calculations

### Summary

Sample MFIs were translated to qualitative (i.e., binary positive/negative) outcomes as described in reference (3) and below. Briefly, for the blood (i.e., serum) samples, empirical training data (in the form of positive and negative controls) were taken as approximate probability models of measurement outcomes for each antigen, conditioned on knowing the class of the underlying sample. Given the training data, we first determined the prevalence *q* (and 95% confidence intervals [CI]) of the test population according to Eq. (4) of Ref. [3]. This estimate was used to adaptively determine a quadratic cutoff boundary that minimizes the classification error *of the test population*. This error is given by $E=q(1-S_e) + (1-q)(1-S_p)$, where the sensitivity $S_e$ and specificity $S_p$ were computed with respect to the training data. The analysis was applied to multidimensional data by treating up to three antigens as distinct axes in a coordinate space. Thus, the classification boundaries were allowed to be high-dimensional surfaces; see **Supplemental Figure 1**. To determine the positive/negative outcomes of the test samples, MFI points falling on one side of the classification surface $S_c$ ($S_{c,+}$), were labeled positive, and those falling on the other ($S_{c,-}$) were labeled negative.

Due to lack of collection method- and population-matched controls, the saliva-based IgG seroprevalence calculations were determined without well-defined positive training data, and alternative control samples from a 2020 Kenyan study were used as a proxy for negative training data. Because these two datasets were expected to have significantly different fractions of positive individuals (which are unknown *a priori*), we estimated the prevalence for both populations using the more advanced analysis of Ref. [3] applicable to *impure training data.* Briefly, we (i) treated the available data as if it were pure; (ii) constructed a family of boundaries that minimize the empirical classification error defined in terms of these populations by varying the corresponding "pseudo-prevalence;" and (iii) solved a resulting system of nonlinear equations that yield the unknown prevalence estimates of the populations. This analysis can be adapted to estimate an analog of 95% CIs; see Ref. [3].

### Determining Qualitative Outcomes for Blood/Serum Samples

Sample MFIs were classified as negative or positive according to the methods of reference (3). As a first step, all data were log-transformed according to the methods of reference [2]. This set the characteristic scale of the data to be of order unity, which is useful for stabilizing all subsequent numerical computations. Next, empirical training data (in the form of positive and negative controls) were taken as approximate probability models of measurement outcomes for each antigen conditioned on the class of the underlying sample. The boundary separating classes were not assumed to be a fixed object, but rather a variable that changes with prevalence to minimize classification error; see reference [2]. Thus, given the training data, the first step in the analysis was to determine the prevalence *q* of the *test population* according to the unbiased, classification-free prevalence estimate of reference [2]. The second step was to determine the cutoff boundary that minimized the total classification error as a function of *q*. Of note, this analysis was applied to *M*-dimensional ($1 \leq M \leq 3$) vectors whose components are log-transformed MFI values associated with a specific antigen. This yields classification boundaries that are *M-1* dimensional surfaces or manifolds; see **Supplemental Figure 1** for a three-dimensional classification boundary example based on RBD, S, and N training data.

<u>Prevalence estimation:</u> To estimate the prevalence $q$ of the test population, the analysis first constructed an *M-1* dimensional quadratic surface *S* that maximized the fractions $Q_p$ and $Q_n$ of positive and negative *training* samples on opposite sides of the manifold, subject to the constraint that $Q_p$=$Q_n$. Refer to the side with more positives (negatives) as $S_+$ ($S_-$). *Note that this S was not used for classification*. Next, let $Q_t$ denote the fraction of *test* samples falling on side $S_+$. The prevalence was then estimated using the equation $q=(Q_t + Q_n - 1)/(Q_p + Q_n - 1)$, which is a mathematically unbiased and converging estimator.

A bound on the variance of the prevalence estimate is given by the formula:

$$\sigma^2 = \frac{(\rho_{max})(1-\rho_{max})}{S(1-2\rho_{max})^2} + \frac{q(1-q)}{S},$$

where $\rho_{max} = 1 - Q_p$ and $S$ is the number of samples. This quantity is an upper bound on the prevalence uncertainty given by the estimator described above; derivation and justification thereof is provided in a forthcoming manuscript. The approximate 95% confidence intervals were then estimated to be $q \pm 2\sigma$. In cases for which the prevalence estimate was 100 %, we instead used the "rule of three", defining the confidence interval to be $\left[1 - \frac{3}{S}, 1\right]$.

<u>Classification:</u> Given a prevalence estimate, the analysis then determined a new, *M-1* dimensional classification surface $S_c$ such that points falling on one side, call it $S_{c,+}$, were labeled positive, and those falling on the other ($S_{c,-}$) were labeled negative. In this work, $S_c$ was defined as the quadratic manifold that minimized the error $E=q(1-S_e) + (1-q)(1-S_p)$, where the sensitivity $S_e$ and specificity $S_p$ were computed with respect to the training data. Note that E depended on $q$.

**Determining Qualitative Outcomes for Saliva Samples**

For saliva data, the classes of the samples in the two available populations (Kenyan samples and US samples) were not known a priori. This means that we had to solve an "unsupervised" problem to determine both the prevalence estimates and classes of each sample.

Because the two populations were sampled at different times in the pandemic, it was reasonable to assume that they had different prevalence outcomes. This allowed us to leverage a novel set of data analysis techniques that can yield exact solutions. We refer the reader to reference [2] for complete details. Briefly, despite the two populations having unknown prevalence estimates, we temporarily assumed that they are pure in that one is treated as corresponding to a "pseudo-prevalence" of $\delta = 0$ and the other is treated as if $\delta = 1$. This pseudo-prevalence is interpreted as the probability that a sample at random from a test population belongs to one of the two impure populations. We then found a family of boundaries that minimized the classification error as a function of the variable pseudo-prevalence. This led to a system of nonlinear equations in terms of the numbers of samples on either side of the classification boundaries.

Ideally, the solution to this system yields a single prevalence estimate for each of the impure populations. However, in practice, two problems arise. First, this system is defined in terms of binomial random variables with a non-zero variance. We thus estimated the 95% confidence range of prevalence estimates as those for which the nonlinear system is solved to within two standard deviations of the random terms of the equation. Second, we can only estimate uncertainties on a finite grid of pseudo-prevalence values. Thus, in cases, where there is only one grid-point satisfies the equations to within uncertainties, we use estimate the uncertainty in prevalence as twice the standard deviation of the corresponding binomial random variable. In cases where prevalence is 100 %, we instead use the “rule of three” estimate.

#### Statistical analysis tools

Statistical calculations and graphs were done in Prism v9.4.1, R v2023.09.1+494, MATLAB R2023a Update 5 (9.14.0.2337262), and SeroNIST Beta Version 0.10, 0.11, and 0.12 (MATLAB scripts based on method described in reference (3)).

**S1 Table.** Self-reported vaccine uptake and pre-existing health conditions among study participants (n=290).

| Category | n (%) |
|---|---|
| **COVID-19 Vaccine** | |
| Yes | 265 (91.4) |
| No | 18 (6.1) |
| Missing | 7 (2.4) |
| **Vaccine Type*** | |
| Moderna | 121 (45.7) |
| Pfizer | 106 (40.0) |
| Johnson & Johnson | 23 (8.6) |
| Other | 15 (5.7) |
| **Flu Vaccine**** | |
| Yes | 230 (79.3) |
| No | 53 (18.3) |
| Missing | 7 (2.4) |
| **Pre-ex. Conditions***** | |
| Hypertension | 32 (11.0) |
| Obesity | 26 (9.0) |
| Diabetes II | 25 (8.6) |
| Asthma | 24 (8.3) |
| Chronic Disease (any) | 21 (7.2) |
| Diabetes I | 9 (3.1) |
| Psych. Condition | 9 (3.1) |
| Cancer | 7 (2.4) |

| | |
|---|---|
| Substance Use Dis. | 6 (2.1) |
| CND | 4 (1.4) |
| Hepatitis | 3 (1.0) |
| Auto/Immunocomp. | 3 (1.0) |
| Alcoholism | 2 (0.7) |
| CKD | 2 (0.7) |
| CLD | 1 (0.3) |
| CRD | 1 (0.3) |
| Dementia | 1 (0.3) |
| Epilepsy | 1 (0.3) |
| None | 168 (57.9) |
| **Smoke/Vape** | |
| No | 241 (83.1) |
| Yes | 39 (13.5) |
| Missing | (3.4) |

*Among those who reported getting the COVID-19 vaccine (n=265), these are the vaccine types received for the first series.
**Self-reported receiving influenza vaccine in the past 5 years.
***Self-reported pre-existing health conditions: A subset of participants reported more than one pre-existing health condition (i.e., percent do not add up to 100%). Psych. conditions: Psychological and mental conditions, Substance Use Dis.: Substance Use Disorder, CND: Chronic Neurological Conditions, Auto/Immunocomp: Autoimmune and immunocompromised conditions, CKD: Chronic kidney disease, CLD: Chronic liver disease, CRD: Chronic respiratory disease.

**S2 Table.** SARS-CoV-2 serum-based *IgG* seroprevalence for each antigen (RBD, S, and N) and combination listed in percent (%) with 95% confidence intervals (95% CI), along with respective predicted classification accuracy (accuracy), sensitivity, and specificity and the CI range. The accuracy, sensitivity, and specificity were calculated based on the training data (i.e., pre-defined positive and negative controls). The associated classification boundaries for each antigen combination were determined adaptively as a function of the study data prevalence to maximize the classification accuracy when the analysis was applied to the study data, while being subject to sensitivity constraints (min. 70% sensitivity).

| | Serum IgG | Accuracy | Sensitivity | Specificity |
|---|---|---|---|---|
| | % (± 95% CI) | % (CI range) | % (CI range) | % (CI range) |
| RBD, S, N | 97.5 (2.4) | 100.0 (96.7, 100.0) | 100.0 (94.0, 100.0) | 100.0 (97.3, 100.0) |
| RBD, S | 99.9 (3.4) | 96.9 (93.0, 99.4) | 98.0 (93.3, 100.0) | 96.3 (92.5, 99.1) |
| S, N | 97.2 (2.0) | 100.0 (96.7, 100) | 100.0 (94.0, 100.0) | 100.0 (97.3, 100.0) |
| RBD, N | 96.5 (2.2) | 100.0 (96.7, 100.0) | 100.0 (94.0, 100.0) | 100.0 (97.3, 100.0) |
| RBD | 96.5 (2.2) | 100.0 (96.7, 100.0) | 100.0 (94.0, 100.0) | 100.0 (97.3, 100.0) |
| S | 97.9 (1.7) | 100.0 (96.7, 100.0) | 100.0 (94.0, 100.0) | 100.0 (97.3, 100.0) |
| N | 49.9 (7.0) | 94.0 (90.2, 97.4) | 86.3 (76.0, 95.1) | 97.5 (94.3, 100.0) |

**S3 Table.** SARS-CoV-2 serum-based *IgA* seroprevalence for each antigen (RBD, S, and N) and combination listed in percent (%) with 95% confidence intervals (95% CI), along with respective predicted classification accuracy (accuracy), sensitivity, and specificity and the CI range. The accuracy, sensitivity, and specificity were calculated based on the training data (i.e., pre-defined positive and negative controls). The associated classification boundaries for each antigen combination were determined adaptively as a function of the study data prevalence to maximize the classification accuracy when the analysis was applied to the study data, while being subject to sensitivity constraints (min. 70% sensitivity).

| | Serum IgA | Accuracy | Sensitivity | Specificity |
|---|---|---|---|---|
| | % (± 95% CI) | % (CI range) | % (CI range) | % (CI range) |
| RBD, S, N | 87.2 (6.4) | 95.1 (90.7, 99.0) | 98.0 (93.2, 100.0) | 92.6 (85.2, 98.3) |
| RBD, S | 83.1 (6.7) | 94.2 (89.3, 98.1) | 95.9 (89.4, 100.0) | 92.6 (84.9, 98.3) |
| S, N | 84.8 (6.6) | 96.1 (91.9, 99.1) | 100.0 (93.9, 100.0) | 92.6 (84.6, 98.3) |
| RBD, N | 39.8 (12.2) | 83.5 (76.2, 90.3) | 89.8 (80.5, 97.7) | 77.8 (66.1, 88.4) |
| RBD | 62.7 (14.5) | 80.8 (73.0, 88.2) | 78.0 (66.0, 88.9) | 83.3 (72.7, 92.9) |
| S | 84.0 (6.7) | 94.2 (89.3, 98.2) | 95.9 (89.2, 100.0) | 92.6 (85.1, 98.3) |
| N | 14.1 (25.5) | 80.6 (73.3, 87.6) | 65.3 (51.9, 78.4) | 94.4 (87.5, 100.0) |

**S4 Table.** SARS-CoV-2 saliva-based IgG seroprevalence for each antigen (RBD, S, and N) and combination in percent (%) and associated uncertainty ranges (uncertainty range, %). The listed seroprevalences do not have respective classification accuracies, sensitivities, and specificities because we calculated the saliva-based seroprevalence assuming impure training data as described in results, methods and reference (3). Hence, without knowing the true classes of the training data we could not estimate the predicted classification accuracies, sensitivities, and specificities. *The N-based saliva seroprevalence resulted in high level of uncertainty due to extensive overlap in MFI between the study sample and control groups (lack of separation in the population-specific outcomes).

| | Saliva IgG |
|---|---|
| | % (uncertainty range,%) |
| RBD, S, N | 100.0 (98.7 - 100.0) |
| RBD, S | 100.0 (98.7 - 100.0) |
| S, N | 96.0 (92.4 - 99.6) |
| RBD, N | 86.9 (81.6 - 96.4) |
| RBD | 86.9 (75.8 - 96.2) |
| S | 96.0 (92.4 - 99.6) |
| N | 48.0 (48.0 - 99.7)* |

**S5 Table.** Coupled antigen type and source.

| **Antigen** | **Virus** | **Source/Company** | **Additional Identifier** |
|---|---|---|---|
| RBD Wuhan (WT) | SARS-CoV-2 | MassBiologics of UMass Chan Medical School | Lot #: 021820 RS 040780 |
| Nucleocapsid Wuhan (WT) | SARS-CoV-2 | MassBiologics of UMass Chan Medical School | Lot #: 060420B RS 063020 |
| Wild-type Full Length Spike Trimer | SARS-CoV-2 | Frederick National Laboratory, Icahn School of Medicine at Mt. Sinai, NCI SeroNet Consortium | SARS-CoV-2 S-2P(15-1213)-T4f-His6 protein, Lot # :P210721.02 |
| HCoV-229E Spike | Endemic coronavirus | Sino Biology | 40605-V08B |
| HCoV-NL63 Spike | Endemic coronavirus | Sino Biology | 40604-V08B |

| | | | |
|---|---|---|---|
| HCoV-OC43 Spike | Endemic coronavirus | Sino Biology | 40607-V08B |
| HCoV-HKU1 Spike | Endemic coronavirus | Sino Biology | 40021-V08H |
| BSA | -- | Sigma Aldrich | A7030-100G |
| RBD alpha | SARS-CoV-2 | MassBiologics of UMass Chan Medical School(4) | |
| RBD beta | SARS-CoV-2 | MassBiologics of UMass Chan Medical School(4) | |
| RBD gamma | SARS-CoV-2 | MassBiologics of UMass Chan Medical School(4) | |
| RBD delta | SARS-CoV-2 | MassBiologics of UMass Chan Medical School(4) | |
| RBD lambda | SARS-CoV-2 | MassBiologics of UMass Chan Medical School(4) | |
| RBD omicron | SARS-CoV-2 | Frederick National Laboratory, Icahn School of Medicine at Mt. Sinai, NCI SeroNet Consortium | TPA-CoV-2-S(318-529)-3C-His8-SBP B.1.1.529, RP1211220.112, Lot #: P211220.02 |

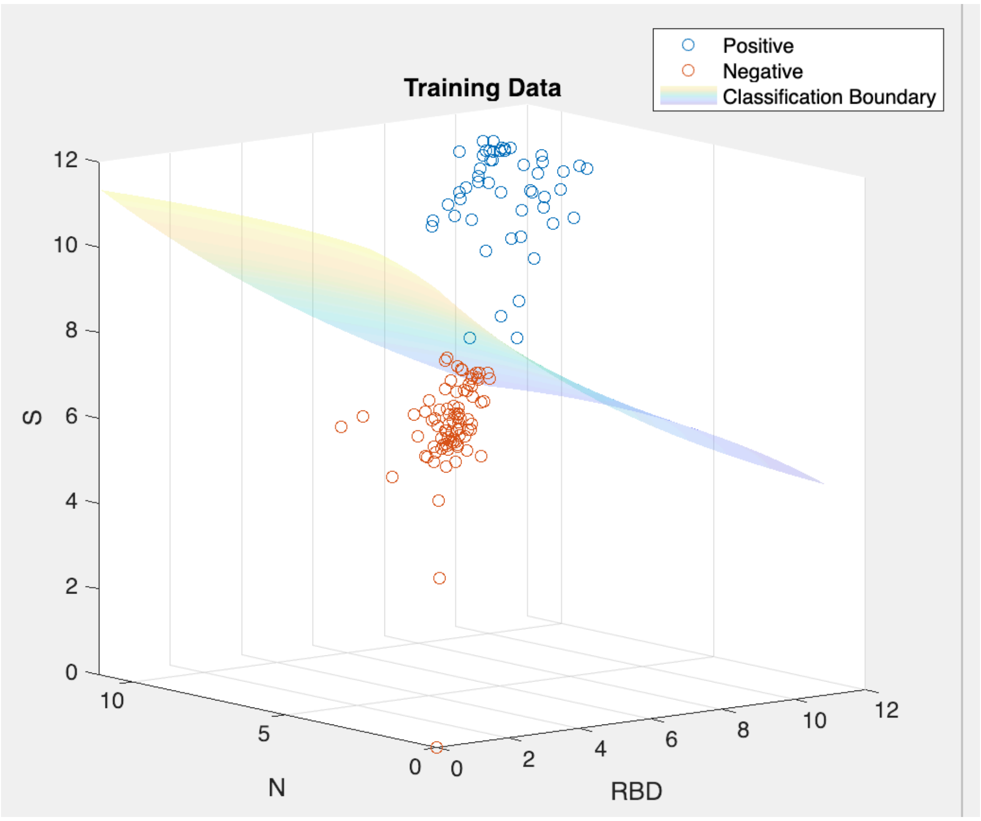

**S1 Fig. Representative graphic of a three-dimensional classification boundary.** The graphic of a three-dimensional (3D) classification boundary graphic was based on training data covering anti-RBD, -S, and -N antibodies from confirmed positive and negative samples. S, Spike. RBD, Receptor Binding Protein. N, Nucleocapsid.

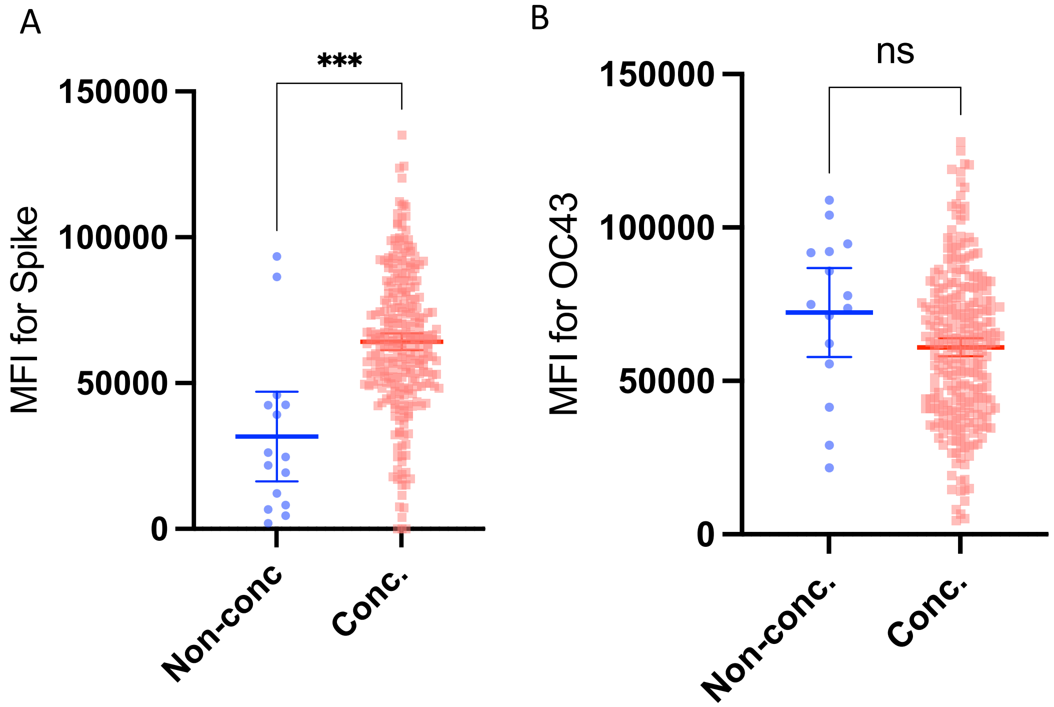

**S2 Fig. Analysis of samples that were non-concordant between the multiplex analysis and the point-of care test.** Comparison of samples that were positive for the multiplex assay and negative for the point-of care test (POC) test across all antigen combinations). (A) The average SARS-CoV-2 spike (S) median fluorescence intensity (MFI) of the non-concordant samples (n=15) was lower compared to the concordant samples (n=265, p=0.0005). (B) The S-based OC43 measurements of the non-concordant samples were not significantly higher than the concordant samples (p=0.12). Hence, the multiplex assay was more sensitive compared to the POC test and but not more likely to be picking up OC43 among the non-concordant samples. The statistical outcomes may be affected by the unequal sample sizes (15 vs 265). MFI, median fluorescence intensity. S, Spike. Ns, non-significant (p>0.05). Non-conc., non-concordant samples (multiplex assay vs. POC test). Conc., concordant samples (sample that had the same qualitative outcome both with the multiplex assay and POC test). POC, point-of-care test. *** = p<0.001, Welch's t-test.

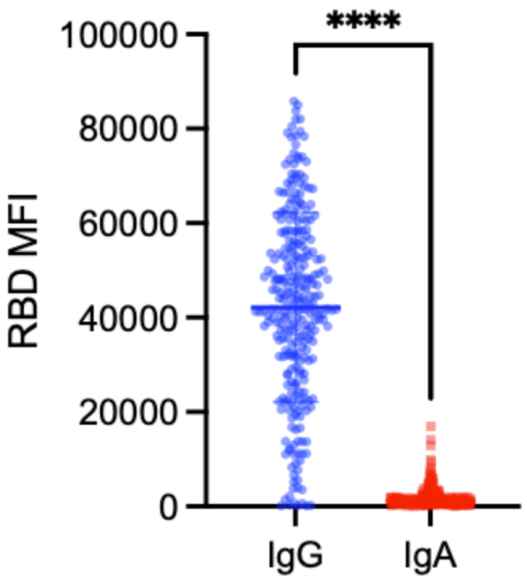

**S3 Fig. Comparison of receptor binding protein-specific serum IgG and IgA outcomes.** Comparison of receptor binding protein (RBD)-specific serum IgG and IgA outcomes as median fluorescence intensity (MFI, mean and standard deviations) among the study participants (mean and standard deviations). **** = p<0.0001, Welch's t-test. MFI, median fluorescence intensity. RBD, Receptor Binding Protein.

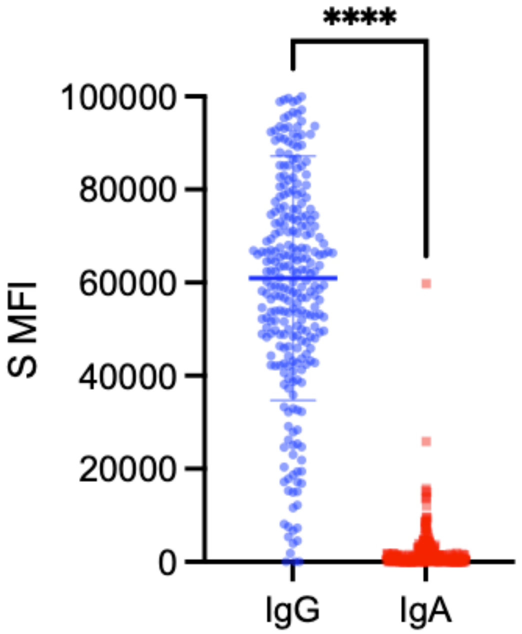

**S4 Fig. Comparison of spike-specific serum IgG and IgA outcomes.** Comparison of spike (S) protein-specific serum IgG and IgA outcomes (MFIs) among the study participants (mean and standard deviations). **** = p<0.0001, Welch's t-test. S, Spike Protein.

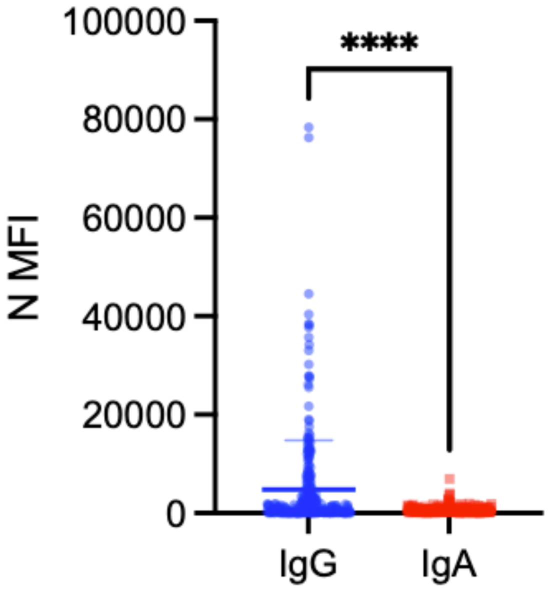

**S5 Fig. Comparison of nucleocapsid-specific serum IgG and IgA outcomes.** Comparison of nucleocapsid (N) protein-specific serum IgG and IgA outcomes (MFIs) among the study participants (mean and standard deviations). **** = $p<0.0001$, Welch's t-test. N, Nucleocapsid Protein.

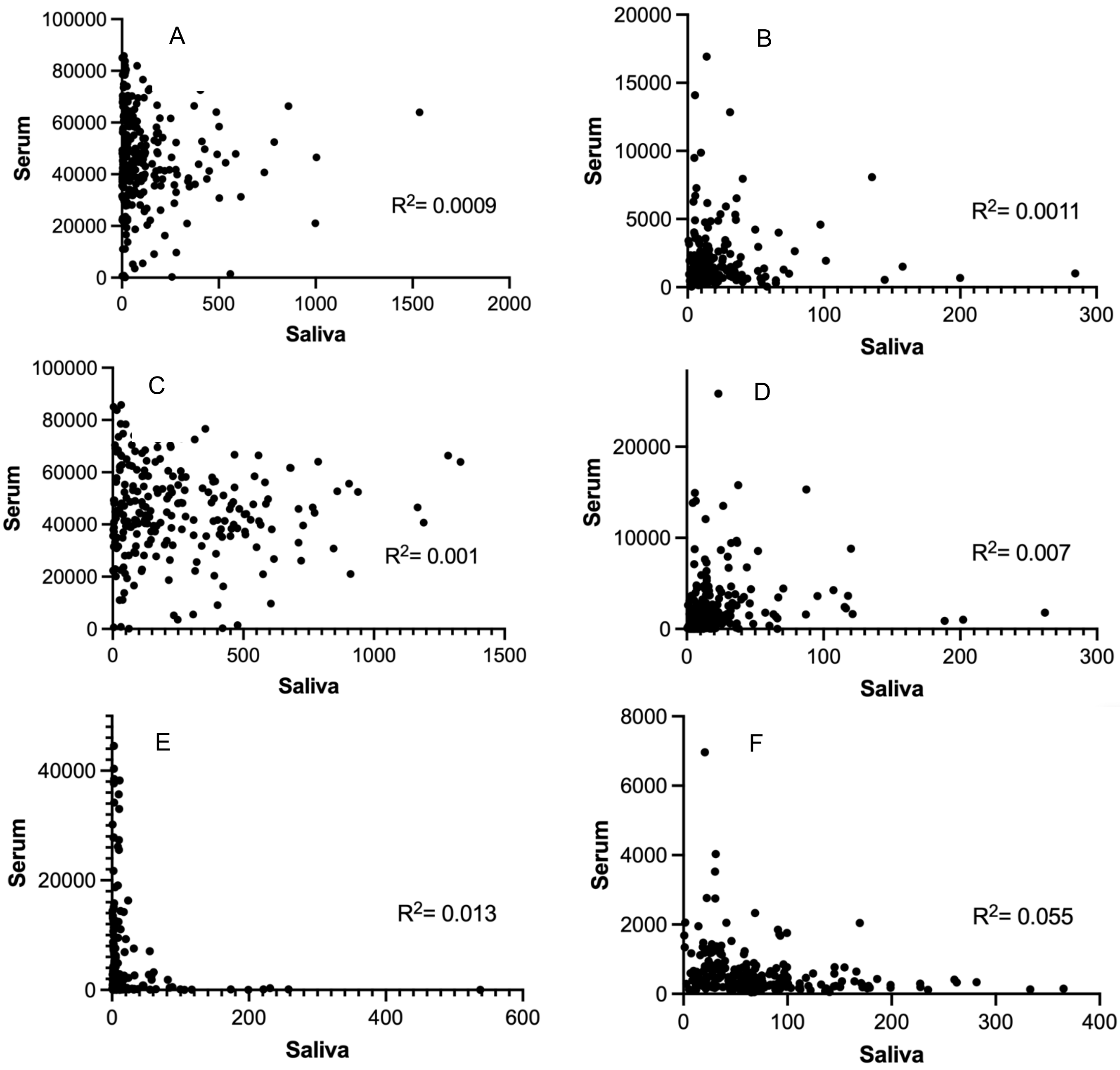

**S6 Fig. Comparison of serum- and saliva-based serological outcomes.** Line up of saliva versus serum comparisons of antigen-specific outcomes (MFI minus BSA for serum and transformed MFI for saliva [antigen- and isotype-specific MFI minus BSA, divided by total Ig, multiplied by 1000]), for anti-SARS-CoV-2 receptor binding domain (RBD; **A, B**), spike (S; **C, D**), and nucleocapsid (N; **E, F**) IgG (left column) and IgA (right column) antibody measurements. The outcomes between serum and saliva did not correlate for any antigen or isotype combination.

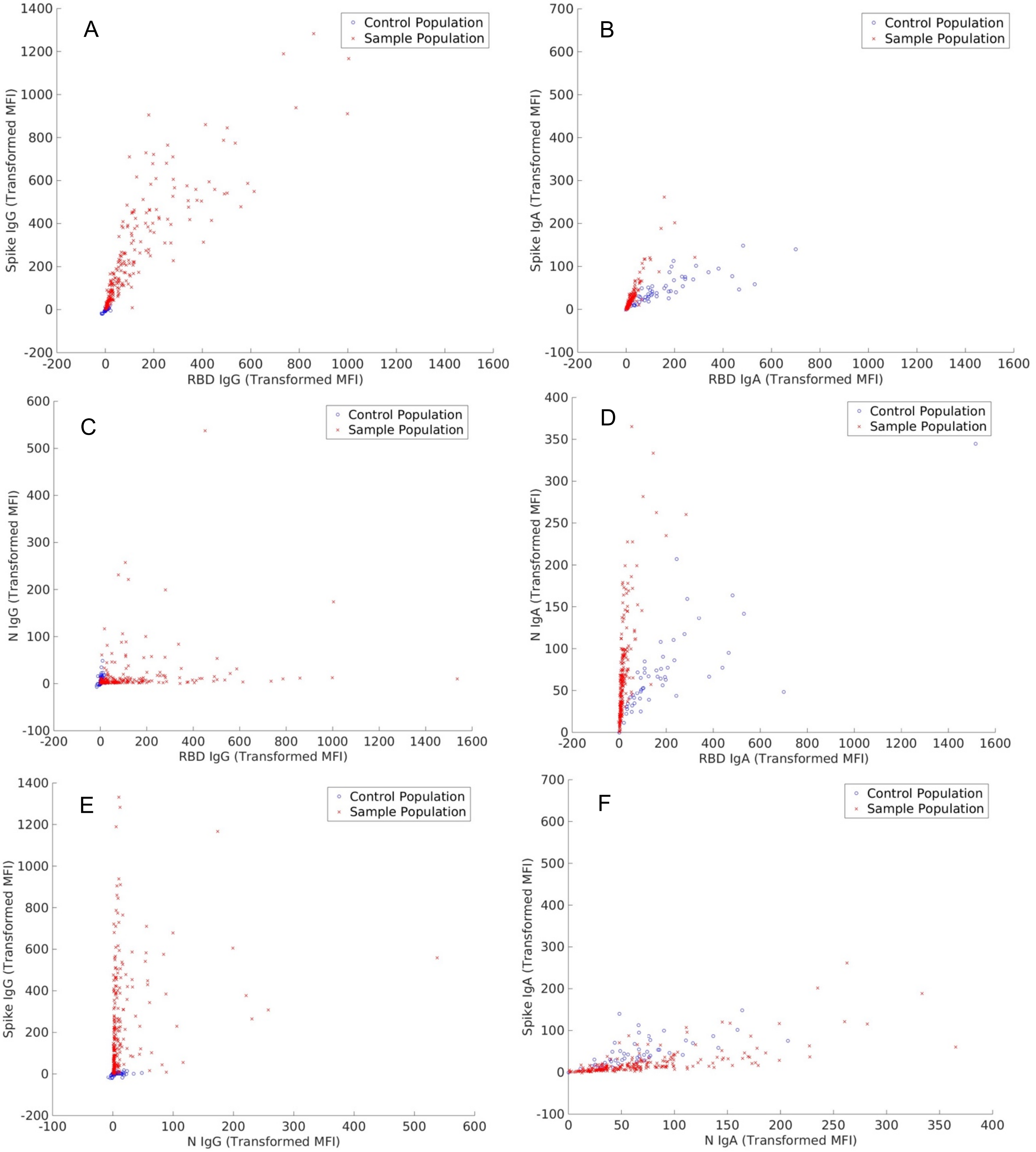

**S7 Fig. Comparison of serological outcomes from study versus control samples.** Comparison of study and control population by line up of saliva-based antigen-specific transformed MFI (antigen- and isotype-specific MFI minus BSA, divided by total Ig, and multiplied by 1000) for IgG (left column) and IgA (right column). For saliva IgG, the control sample population (sample collection method-matched samples from Kenya) always clusters in low MFI area and separate well from the study sample population (**A, C, E**), whereas for IgA the outcomes/MFIs from the control sample population overlap significantly with the study sample population for at least one antigen (**B, D, F**) and score higher maximum MFIs for RBD-specific outcomes (**B, D**). Hence, no saliva IgA percent seroprevalences could be calculated for the study samples based on these controls. S, Spike. RBD, Receptor Binding Protein. N, Nucleocapsid.

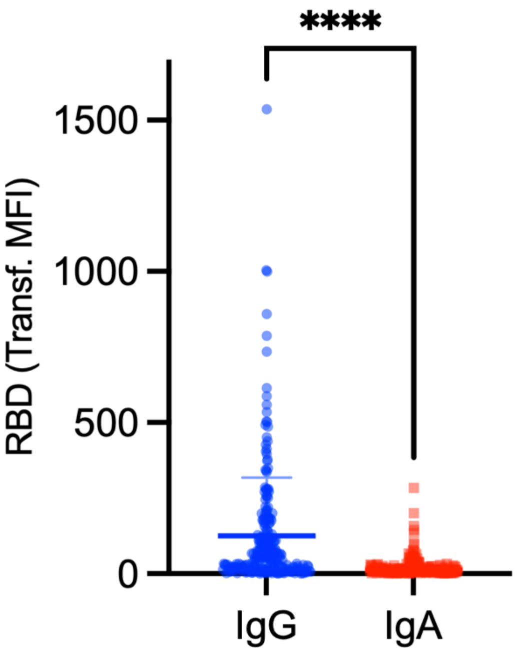

**S8 Fig. Comparison of receptor binding protein-specific saliva IgG and IgA serological outcomes.** Comparison of RBD-specific saliva IgG and IgA outcomes (transformed MFI = [raw MFI/total Ig]*1000) among the study participants (mean and standard deviations). **** = p<0.0001, Welch's t-test. RBD, Receptor Binding Protein.

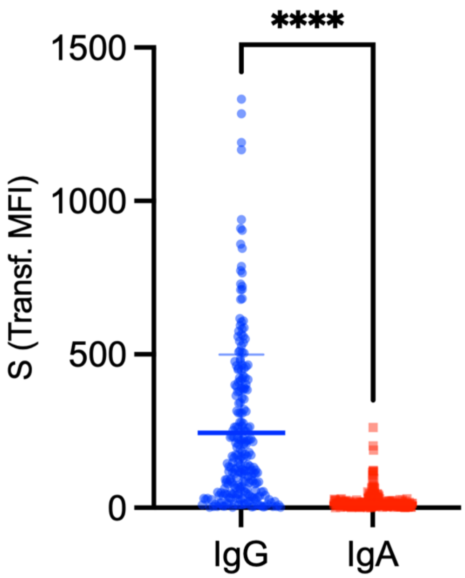

**S9 Fig. Comparison of spike protein-specific saliva IgG and IgA serological outcomes.** Comparison of S-specific saliva IgG and IgA outcomes (transformed MFI = [raw MFI/total Ig]*1000) among the study participants (mean and standard deviations). **** = p<0.0001, Welch's t-test. S, Spike.

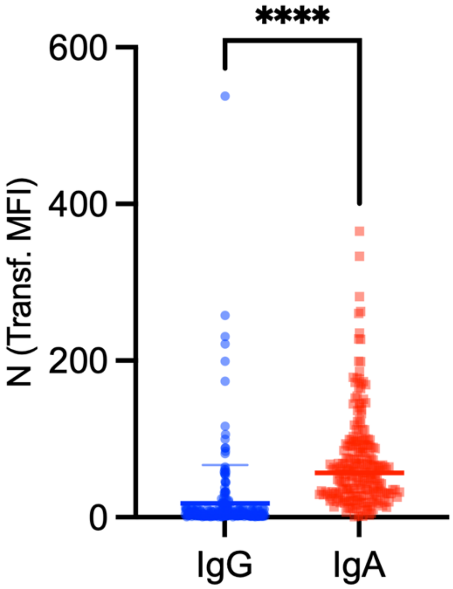

**S10 Fig. Comparison of nucleocapsid protein-specific saliva IgG and IgA serological outcomes.** Comparison of nucleocapsid (N)-specific saliva IgG and IgA outcomes (transformed MFI = [raw MFI/total Ig]*1000) among the study participants (mean and standard deviations). **** = p<0.0001, Welch's t-test. For N-specific outcomes in saliva, the IgA reads are higher compared to IgG. Whereas for RBD and S, the IgG reads in saliva are higher. Overall, the N-specific saliva IgG and IgA outcomes (transformed MFI) are lower than for RBD and S (i.e., lower overall MFI, see y-axis comparison between Supplemental Figures 8, 9, and 10). N, Nucleocapsid.

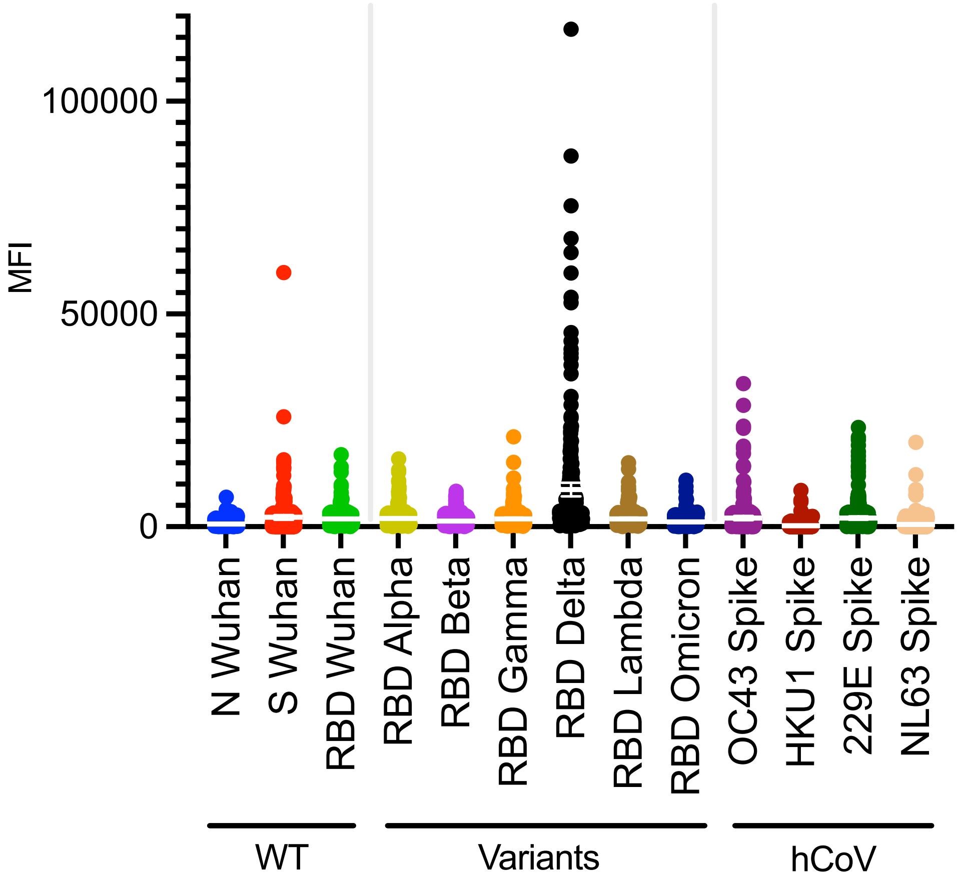

**S11 Fig. Serum IgG outcome distribution in linear scale.** Linear scale dot plot with means and 95% confidence intervals (CI) of antigen-specific antibody measurements (MFI minus BSA and normalized across plates) for serum IgG SARS-CoV-2 and variants, along with human endemic coronaviruses OC43, HKU1, NL63, 229E. MFI, median fluorescence intensity. N, Nucleocapsid. S, Spike. RBD, Receptor Binding Protein. hCoV, human endemic coronaviruses. WT, wild-type (Wuhan).
